# Complement C3 vs C5 inhibition in severe COVID-19: early clinical findings reveal differential biological efficacy

**DOI:** 10.1101/2020.08.17.20174474

**Authors:** Dimitrios C. Mastellos, Bruno G. P. Pires da Silva, Benedito A. L. Fonseca, Natasha P. Fonseca, Maria A. Martins, Sara Mastaglio, Annalisa Ruggeri, Marina Sironi, Peter Radermacher, Panagiotis Skendros, Konstantinos Ritis, Ilenia Manfra, Simona Iacobelli, Markus Huber-Lang, Bo Nilsson, Despina Yancopoulou, E. Sander Connolly, Cecilia Garlanda, Fabio Ciceri, Antonio M. Risitano, Rodrigo T. Calado, John D. Lambris

**Author notes:** these authors contributed equally (Dimitrios C. Mastellos, Bruno G. P. Pires da Silva). corresponding author, (JDL).

## Abstract

Growing clinical evidence has implicated complement as a pivotal driver of COVID-19 immunopathology. Deregulated complement activation may fuel cytokine-driven hyper-inflammation, thrombotic microangiopathy and NET-driven immunothrombosis, thereby leading to multi-organ failure. Complement therapeutics have gained traction as candidate drugs for countering the detrimental consequences of SARS-CoV-2 infection. Whether blockade of terminal complement effectors (C5, C5a, or C5aR1) can elicit similar outcomes to upstream intervention at the level of C3 remains debated. Here we have compared the clinical efficacy of the C5-targeting mAb eculizumab with that of the compstatin-based C3-targeted drug candidate AMY-101 in small independent cohorts of severe, mainly non-intubated COVID-19 patients. Our exploratory study indicates that therapeutic complement inhibition abrogates COVID-19 hyper-inflammation. Both C3 and C5 inhibitors elicit a robust anti-inflammatory response, reflected by a steep decline in CRP and IL-6 levels, associated with marked lung function improvement and resolution of SARS-CoV-2-associated ARDS. C3 inhibition afforded broader therapeutic control in COVID19 patients by attenuating both C3a and sC5b-9 generation and preventing FB consumption. This broader inhibitory profile of anti-C3 treatment was associated with a more robust decline of neutrophil counts, a greater decline of median LDH levels and more prominent lymphocyte recovery within the first 7 days of treatment. These early clinical results offer important insight into the differential mechanistic basis and underlying biology of C3 and C5 inhibition in COVID-19. They point to a broader pathogenic involvement of C3-mediated pathways and set the stage for larger prospective trials that will benchmark these complement-targeting agents in COVID-19.

## Introduction

As the COVID-19 pandemic sweeps through the globe with an increasing death toll, pressing questions about its intricate immunobiology arise, pointing at the same time to the urgent need for effective therapeutic measures to combat the detrimental systemic consequences of this disease ^1, 2, 3^. Although initially perceived as a disease that exclusively targets the respiratory tract of individuals infected by SARS-CoV-2, COVID-19 has evolved into a complex, multi-organ pathology with a plethora of thomboinflammatory manifestations in key vital organs, including the lungs, heart, liver, kidney, and brain ^2^. The common denominator driving pathology in these organs appears to be an extensive and deregulated activation of innate immune pathways that leads to massive monocyte and neutrophil infiltration into infected tissues and to a disseminated thromboinflammatory response of the microvascular endothelium (thrombotic microangiopathy) ^4, 5, 6^. This derailed inflammatory response, marked by a systemic increase in proinflammatory cytokines (known as cytokine storm) reflects a maladaptive host immune response to SARS-CoV-2 that is instigated by pathogen recognition systems, such as the complement cascade, which becomes overwhelmingly active in their attempt to thwart the infectious agent ^5, 7^.

Complement dysregulation is emerging as a key driver of COVID-19 hyper-inflammation, immunothrombosis and microvascular endothelial injury ^5, 7, 8^. Systemic complement activation, predominantly via the lectin pathway (LP) and classical pathway (CP), is closely correlated with microvascular injury, platelet-neutrophil activation, and a NET-dependent, Tissue Factor (TF)-driven hypercoagulable phenotype that disseminates through the vascular bed of multiple organs ^8^. Mounting clinical data have implicated deregulated complement and coagulation pathways as risk factors for adverse outcomes in COVID-19 ^9^. Increased C5a and sC5b-9 levels and prominent activation of the C5a-C5aR1 axis have been reported both in the infected lungs and systemically ^10,11,12^. While these studies have propelled the clinical evaluation of terminal pathway therapeutics (anti-C5, C5a/C5aR1 blockade), key C3-mediated processes that fuel monocyte/neutrophil-driven inflammatory damage, cytokine responses and TF-driven thrombophilia in COVID-19 remain operative ^8^. These include upstream C3 convertase activity, leading to C3b-opsonization and AP amplification via any of the triggering routes that SARS-CoV-2 infection engages.

Earlier studies in animal models of SARS-CoV infection underscored the pivotal role of C3 activation in the pathogenesis of SARS-CoV associated ARDS ^13^. Given that C3 activation is the convergence point of all complement pathways, we hypothesized that C3 targeting may afford broader and more comprehensive therapeutic coverage in COVID-19-associated ARDS ^5^.

Here we have performed a comparative assessment of key clinical and biochemical correlates in two small COVID-19 patient cohorts with SARS-CoV-2 associated ARDS treated either with the C3-based therapeutic AMY-101 (Amyndas) or with the C5-targeting mAb eculizumab (Soliris, Alexion). Patients received AMY-101 within a compassionate use program, and eculizumab within a prospective phase I/II single arm clinical trial. Eculizumab is a clinically approved anti-C5 mAb that targets exclusively the terminal (lytic) pathway and its downstream effectors (C5a and MAC) ^14^. AMY-101 is a C3-targeted drug candidate based on third-generation compstatins, a family of cyclic peptides that bind C3 and prevent its activation by C3 convertases. AMY-101 and earlier compstatin analogs are currently in Phase II/III development, having shown safety in trials of chronic C3 intervention ^15^.

While we acknowledge the inherent difficulty of testing these agents against a heterogeneous clinical background, especially in terms of reconciling disparate supportive measures or patient demographics, we have pursued a thorough investigation of the dynamic profile of changes in key disease markers during treatment with C3 and C5 inhibitors. This exploratory study has revealed both common and distinct clinical correlates and provides important insight into the underlying biology of complement inhibition in severe COVID-19.

## Materials and methods

### Patient cohorts, drug intervention and supportive treatment

In our exploratory study, AMY-101 was administered as a continuous IV infusion at a dose of 5mg/kg/daily to three (3) severe COVID-19 patients with ARDS under a compassionate use program program (CUP) in San Raffaele Hospital, Milan, Italy (Approval March 26^th^ 2020 by Ethical Committee of “Istituto Nazionale per le Malattie Infettive Lazzaro Spallanzani I.R.C.C.S.”, Parere N. 35 del Registro delle Sperimentazioni”). Eculizumab was administered intravenously once a week (1-3 doses of 900 mg) to ten (10) consecutive COVID-19 patients enrolled in a phase I/II single arm clinical trial (http://www.ensaiosclinicos.gov.br, RBR-876qb5) at the University Hospital, University of São Paulo, Ribeirão Preto School of Medicine, Ribeirão Preto (Brazil). Inclusion criteria were age 18-80 years and severe or critical COVID-19 (Wu Z, McGoogan JM. JAMA 2020) confirmed by a positive RT-PCR in the nasopharyngeal swab or tracheal lavage. All patients required oxygen support before treatment initiation; three eculizumab-treated patients were subjected to mechanical ventilation during therapy. Patient demographics and clinical characteristics for both cohorts are presented in Table S1 (supplementary files). All COVID-19 patients received supportive care during anti-complement therapy including anticoagulants and broad-spectrum antibiotics (see Table S1). The Ecu-cohort also received concomitant treatment with corticosteroids according to physician’s assessment. All ecu-patients received penicillin as prophylaxis during treatment.

### Complement activity assay (APH50)

Complement hemolytic activity via the alternative pathway (APH50) was monitored in all patient samples as previously described ^16^. In brief, AP-mediated hemolytic activity was measured by the lysis of rabbit erythrocytes in the presence of patient sera dosed with the C3 and C5 inhibitors. The degree of hemolysis was determined by spectrophotometric analysis of supernatants after centrifugation.

### Complement protein levels and C3/C5 activation fragments

C3, C4, and FB levels were determined by nephelometry in patient plasma samples using an IMMAGE 800 protein chemistry analyzer (Beckman Coulter) C3dg levels were measured by nephelometry following PEG precipitation of patient plasma (11 % w/v) and subsequent incubation with an anti-C3d antibody (Dako). C3a and sC5b-9 levels were measured in EDTA-plasma collected from the patients at predetermined time points by human C3a- and C5b-9-specific ELISAs according to the manufacturer’s instructions (Quidel). sC5b-9 levels were also determined using a modified ELISA method described in ^17^.

To enable comparison of C5b-9 measurements derived from different laboratories and to normalize values generated from C5b-9 ELISA-based assays with different dynamic ranges of detection, we plotted the fold change of C5b-9 levels over baseline values for each patient/cohort.

### Assay for IL-6

IL-6 levels were quantified in patient EDTA-plasma using an ELISA automated immunoassay (R&D Systems) following manufacturer’s instructions.

### Monitoring of AMY-101 levels in patient plasma

Quantitation of AMY-101 was performed in patient EDTA-plasma samples collected at predetermined time points during treatment by UPLC-ESI-MS as described previously ^18^.

### Assays for procoagulant markers

Thrombin-antithrombin complex (TAT) levels were quantified in patient EDTA-plasma using Enzygnost (TAT micro; Behringwerke, Marburg, Germany) as previously described ^17^.

### Statistical analysis

In view of the small sample size no formal comparisons for statistical significance were performed between the two patient cohorts. However, statistical analysis between two data sets (i.e., treatments, days) within the same group was performed using the unpaired (two-tailed) student’s t–test (Prism, GraphPad v 8.0). The level of statistical significance was set to 0.05. Data are presented as mean values +/- standard deviation (SD).

## Results and discussion

Complement intervention has emerged as a promising strategy for ameliorating COVID-19 thromboinflammation. Consistent with this notion, compassionate treatment of a severe COVID-19 patient with the C3 therapeutic AMY-101 abrogated the hyper-inflammatory phenotype associated with SARS-CoV-2, leading to respiratory improvement and resolution of ARDS ^19^. Other reports have indicated that C5 inhibition or downstream C5a blockade may also benefit COVID-19 patients ^20, 11, 21, 22^. While several complement therapeutics are advancing through the biopharma pipeline as potential COVID-19 anti-inflammatories, there has been no attempt to dissect the mechanistic basis of complement inhibition in COVID-19 or benchmark the efficacy of discrete anti-complement agents in COVID-19 patients.

Whether proximal (C3-based) or terminal (C5-based) complement inhibition may afford broader and sustained therapeutic effects in these patients remains the subject of ongoing investigation. Our exploratory study has assessed C3 and C5 inhibition in a series of severe COVID-19 patients, providing early evidence for a differential clinical response and drug efficacy that likely reflects distinct features of the underlying biology of C3 and C5 inhibition.

In the AMY-101 CUP, three patients were recruited from April 10^th^ to May 21^st^, 2020. Treatment was initiated in patients fulfilling the following criteria: COVID-19, diagnosed with qRT-PCR and chest X-ray and/or CT scan; ARDS, defined as acute-onset respiratory failure with bilateral infiltrates on chest radiograph or CT scan, hypoxemia as defined by a PaO_2_:FiO_2_ ratio ≤ 300 mm Hg with a positive end-expiratory pressure (PEEP) of at least 5 cm of H_2_O, and no evidence of left atrial hypertension to rule out cardiogenic edema; hyper-inflammation, defined as elevation of serum inflammation markers C-reactive protein (CRP ≥ 100 mg/L) and/or ferritin (≥ 900 ng/ml) (exclusion criteria listed in supplemental data).

In the eculizumab trial, patients meeting inclusion criteria were recruited from May 25^th^ to June 30^th^, 2020. A total of 12 patients met criteria, but two refused to participate. A total of 10 patients gave written informed consent and were enrolled during this period of time. Two patients who were on mechanical ventilation before enrollment died of mechanical ventilation-associated pneumonia on days +19 (UPN1) and +18 (UPN9). The other eight patients showed clinical improvement, were discharged, and are alive until last follow-up on August 12^th^, 2020. One patient (UPN5) was intubated within 12 hours after the initial dose, required mechanical ventilation until day +9, and was discharged on day +17. All other patients did not require mechanical ventilation, but oxygen supplementation (nasal catheter, high flow oxygen mask), and were discharged on days +2 to +27 (median, 10 days). Three patients received one dose, three patients received two doses, and four patients received three doses of eculizumab. Eculizumab administration was halted when patients showed significant clinical improvement not requiring oxygen supplementation. Eight out of ten patients were alive and discharged at day +28. No severe adverse event (CTCAE grade III or IV) attributable to the drug was observed. Other severe adverse events are described in Table S2 (supplemental data).

## Profiles of clinical response

### Impact of complement inhibitors on markers of COVID-19-associated tissue injury and inflammation

Both C3 and C5 inhibition elicited a robust anti-inflammatory response in COVID-19 patients marked by a rapid decline of C-reactive protein (CRP) levels that led to normalization with 6-8 days after treatment initiation. (Fig 1, panel A). This rapid anti-inflammatory response also was reflected in a reciprocal decrease of IL-6 levels in AMY-101- and Ecu-treated patients that led to an almost 50% reduction of baseline values within 48 h from the start of treatment (Fig 1, panel C). The pronounced tissue protective and anti-inflammatory effect of complement inhibition in COVID-19 patients was indicated by a significant reduction of LDH levels in both cohorts. Of note, AMY-101 treatment correlated with a steeper decline in LDH levels as compared to baseline values, in the first 7 days of treatment (Fig 1). AMY-101 led to a 48.2% decrease of median LDH levels compared to 37.6% in non-intubated eculizumab-treated patients (n=7).

**Figure 1:**
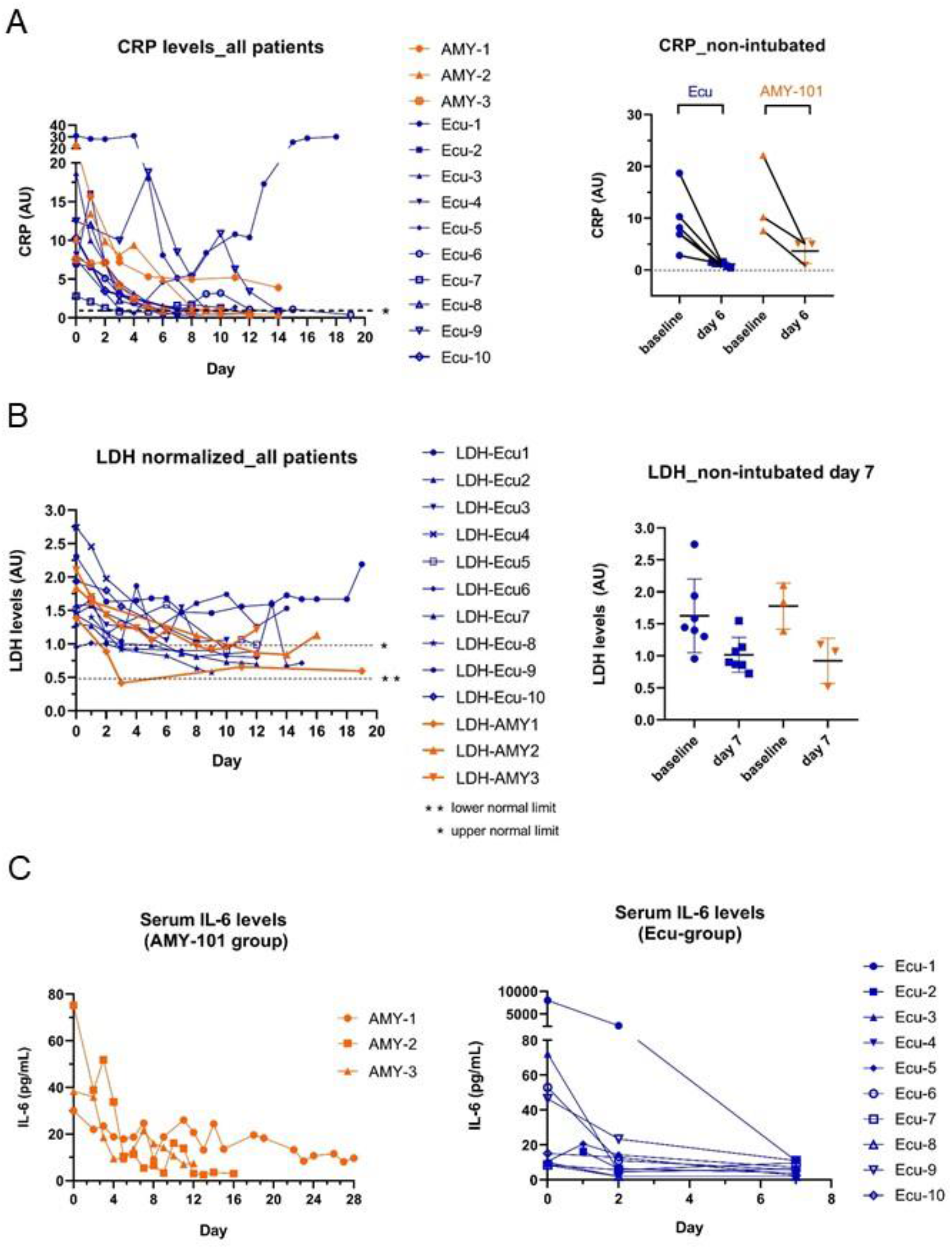
Markers of inflammation and tissue injury in severe COVID-19 patients treated with C3 and C5 inhibitors. Graphs on the left column (panels A, B) represent the longitudinal change of inflammatory and tissue injury-related biomarkers in all COVID-19 patients dosed with the C3 (AMY-101) or C5-targeted inhibitor (eculizumab). These graphs include the three mechanically ventilated Ecu-patients (Ecu-patients No 1, 3, 9). To normalize for disease severity and exclude potential confounding factors from our analysis, the graphs on the right column represent the differential change of these markers in the non-ventilated patients of both cohorts from baseline to days 6-7. Panel A, Change of CRP levels in both patient cohorts; CRP values are expressed as fold change over the upper normal limit of each patient cohort. Panel B, Change of LDH levels in both patient cohorts; LDH values are expressed as fold change over the upper normal limit of each patient cohort. Panel C shows the consistent decrease of serum IL-6 levels in both patient cohorts. The plots illustrating the dynamic profiles of all biomarkers and all individual data points per each patient group are colour-coded (orange: AMY-101-treated, dark blue: Eculizumab-treated).

### Impact on blood cell counts

Elevated neutrophil counts have been correlated with increased disease severity and poor prognosis in COVID-19 patients ^23^. Strikingly, while AMY-101-treated patients exhibited a fast and consistent decline of blood neutrophils numbers, starting as early as 24 h after the initiation of drug infusion (Fig. 2, panel A), Ecu-patients showed persistently elevated neutrophil counts throughout the treatment. The return of neutrophil numbers to normal levels was significantly delayed in Ecu-patients, as shown in Fig. 2 panel A. On day 7, AMY-101-treated patients showed a 51.1% decline in neutrophil counts (baseline ANC=8.6±2.1 cells×10^9^/L, mean ANC value on day 7=4.46±0.58 cells×10^9^/L), while non-intubated Ecu-patients showed no significant change over baseline values (baseline mean ANC=6.52±2.8 cells×10^9^/L, mean ANC value on day 7=7.35±2.8 cells×10^9^/L).

**Figure 2:**
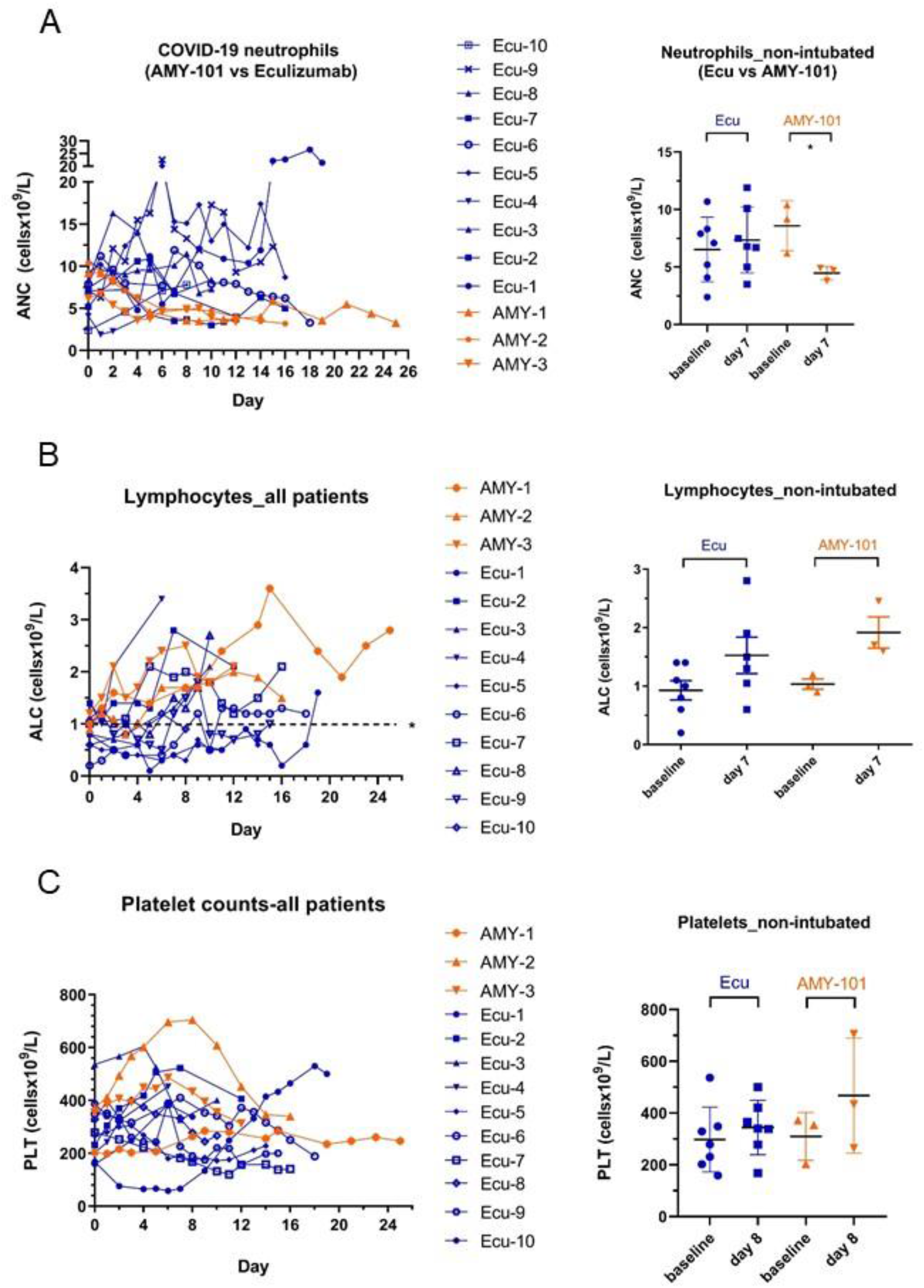
Blood cell monitoring during therapeutic complement inhibition in severe COVID-19. Graphs on the left column (panels A, B and C) represent the longitudinal change of blood cell counts in all COVID-19 patients dosed with the C3 (AMY-101) or C5-targeted inhibitor (eculizumab). These graphs also include the three mechanically ventilated Ecu-patients (Ecu-patients No 1, 3, 9). To normalize for disease severity and exclude potential confounding factors from our analysis, the graphs on the right column represent the differential change of these cell counts in the non-ventilated patients of both cohorts from baseline to days 7 or 8 after the start of drug dosing. *Panel A*, Change of peripheral blood neutrophil numbers in both patient cohorts; neutrophil numbers are expressed as absolute neutrophil counts (ANC, cells×10^9^/L). *Panel B*, Change of peripheral blood lymphocyte numbers in both patient cohorts; lymphocyte numbers are expressed as absolute lymphocyte counts (ALC, cells×10^9^/L). The dotted line represents the upper normal limit of lymphocyte counts in the circulation of healthy individuals. *Panel C* shows the longitudinal change of platelet counts in both patient cohorts. The plots illustrating the dynamic profiles of all biomarkers and all individual data points per each patient group are colour-coded (orange: AMY-101-treated, dark blue: Eculizumab-treated).

One of the cardinal features of COVID-19 is the presence of low lymphocyte counts in severe patients (lymphopenia) ^1^. Lymphopenia on admission is a risk factor associated with a poor prognosis of COVID-19 patients ^24^. In our study, complement inhibition effectively reversed COVID-19 associated lymphopenia, leading to recovery of blood lymphocyte numbers over the course of treatment. Of note, the rate of lymphocyte recovery in the AMY-101 group was faster, with a more prominent increase of mean lymphocyte numbers by day 7 from the start of dosing (AMY-101 group: 85.8% increase of mean ALC, Ecu-group: 65% increase of mean ALC) (Fig.2 panel B). This probably implies a more rapid reversal of the blunted adaptive cellular immune response described in severe COVID-19 patients ^25^.

### Impact on markers of COVID-19 coagulopathy

Given the emerging role of complement dysregulation in COVID-19 immunothrombosis and the presence of thrombocytopenia in severe COVID-19 cases ^8, 26, 23^, we next investigated the impact of complement inhibition on platelet counts and on distinct markers of coagulopathy. C3 inhibition resulted in a steeper transient increase of platelet numbers in COVID19 patients with a trend towards a greater increase in platelet counts between baseline (day 0) and day +8 in the AMY-101 cohort. While this finding indicates a likely more pronounced beneficial effect of C3 inhibition on platelet consumption early during the treatment, C5 blockade was also associated with a transient, albeit more moderate, increase in platelet counts during the same time window (Fig 2, panel C). Signifying a broader downregulation of procoagulant and fibrinolytic responses during complement interception, both D-dimer levels and Thrombin-antithrombin (TAT) complexes were markedly decreased within the 7 first days of treatment in the presence of both inhibitors (supplementary data).

### Lung respiratory function

The robust anti-inflammatory profile and impact of both complement inhibitors on markers of COVID-19 coagulopathy was readily reflected in a marked improvement of lung respiratory function in all non-intubated patients. This improvement culminated in full resolution of ARDS, amelioration of SARS-CoV-2-associated bilateral interstitial pneumonia and weaning off oxygen support in 10-13 days following the start of therapy (average "time to no O_2_ support” for ecu-patients:10.5 days, ranging between 1-27 days; average time for AMY-101-patients: 13.3 days, ranging between 9-18 days) (Fig. 3).

**Figure 3:**
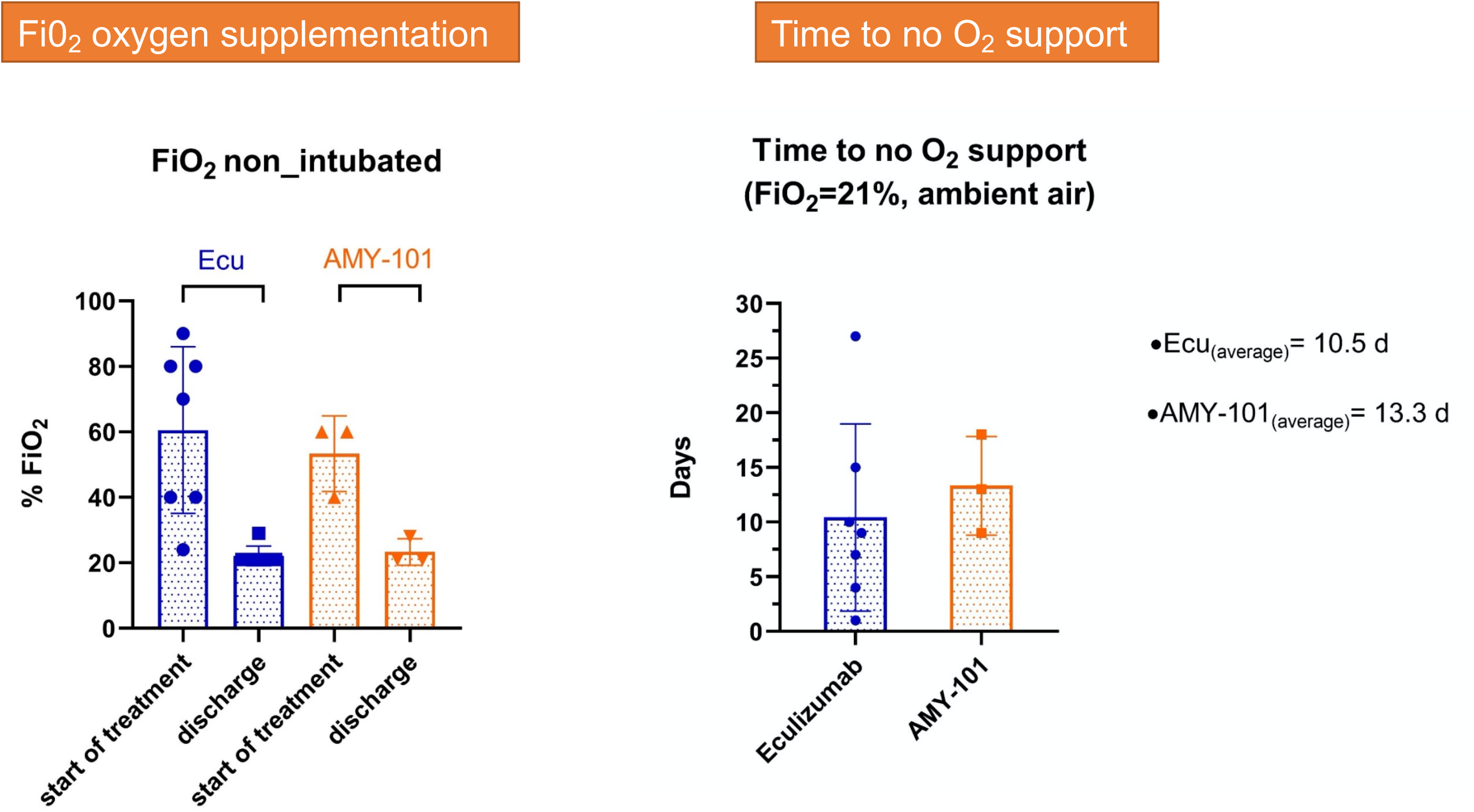
Clinical improvement of lung respiratory function and resolution of SARS-COV-2 associated ARDS. Improvement of lung function in both patient cohorts was monitored as a function of the need for oxygen supplementation (expressed as % FiO_2_; % pf Fraction of Inspired Oxygen in Ventimask). The left graph represents the fraction of patients within each group that were weaned off oxygen support (by breathing in ambient air conditions, or %21 FiO_2_). Bars denote the baseline FiO_2_ values and corresponding values at patient discharge. The right graph illustrates the average time (in days) required for each patient to achieve disengagement from oxygen support (expressed as ‘time to no O_2_ support’). Individual data points and bars are colour-coded according to treatment (Eculizumab, dark blue; AMY-101, orange). Changes are expressed as mean %FiO_2_ values ±SD.

#### Dissecting the *in vivo* biological efficacy of C3 vs C5-targeted inhibition in COVID-19

We next sought to determine whether the qualitative traits suggesting an improved clinical response of AMY-101 over eculizumab were rooted in the different mechanistic basis of complement inhibition and the distinct *in vivo* potency of each inhibitor. To this end, we performed a comparative study of *in vivo* markers of complement activity in each patient cohort. As shown in Fig. 4, C3a levels were significantly attenuated in COVID-19 patients treated with AMY-101, consistent with effective blockage of C3 activity (Fig. 4, panel A). C3a levels dropped sharply from baseline to day +2 (76.5% decline) and remained low throughout the treatment (data shown only until day 7 for comparison with the ecu-group). In contrast, persistently high levels of C3a were detected in the plasma of all eculizumab-treated patients from baseline through day 7 (Fig. 4, panel A), consistent with the notion that eculizumab cannot interfere with upstream C3 activation and C3a release.

**Figure 4:**
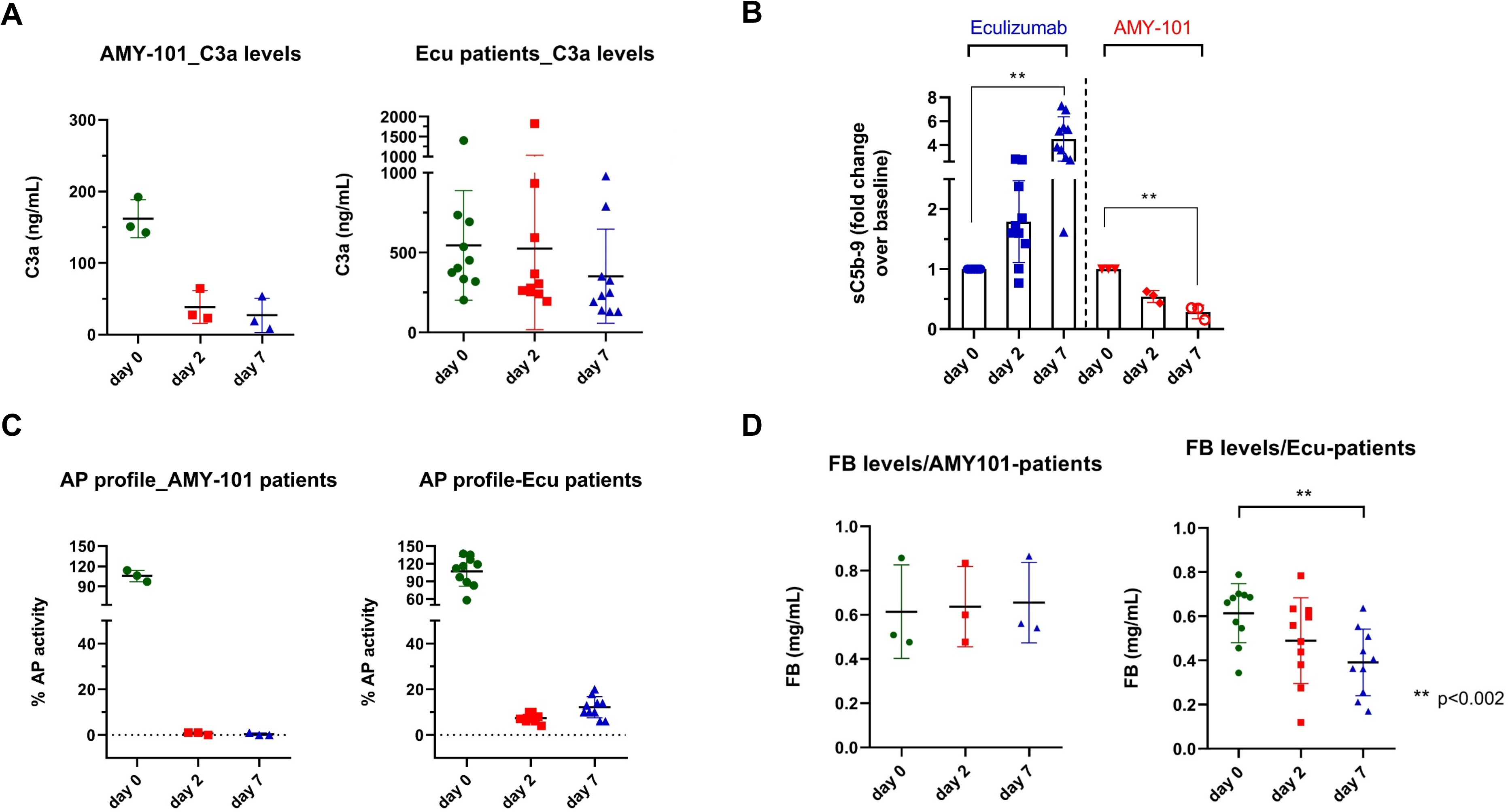
In vivo biological efficacy of C3 vs C5 inhibition in COVID-19 - biomarkers of complement activity. (Panel A): Plasma C3a levels, as a marker of ongoing C3 activation, in patients treated with AMY-101 or eculizumab on days 0 (baseline), 2 and 7 following initiation of treatment. C3a values were quantified in EDTA-plasma samples by ELISA. *Panel B:* Plasma levels of sC5b-9 complexes, as a measure of ongoing terminal pathway activity (C5 activation) in patients treated with AMY-101 or eculizumab. Values for each patient are expressed as fold change over baseline (day 0). *Panel C:* Profiles of AP activity following C3 and C5 inhibition in COVID-19 patients. % AP activity was expressed as the % hemolytic activity of patient sera dosed with each inhibitor using ex vivo AP-mediated complement hemolytic assays (APH50). Panel D: Plasma levels of factor B (FB) in both patient cohorts during treatment with complement inhibitors (days 0-2-7). Total FB was measured in patient plasma by nephelometry using an IMMAGE 800 protein chemistry analyzer. Statistical analysis and comparisons within each group were performed with unpaired, two-tailed student’s T test; ** denotes p <0.002

Treatment with eculizumab was associated with a dissimilar/divergent pattern of C5 blockade in COVID-19 patients. While sC5b-9 levels remained close to baseline values until day 2 (albeit with an upward trend), a significant rebound of sC5b-9 levels was observed on day 7 for most patients under eculizumab treatment (i.e., an almost 5-fold increase over baseline values) (Fig. 4, panel B). This rebound might indicate a breakthrough in C5 inhibition, which could be due to suboptimal C5 blockade associated with overt complement activation (pharmacodynamic PD) and/or insufficient dosing of the drug. This observation follows up on recent reports indicating suboptimal C5 blockade in COVID-19 patients treated with eculizumab, under a similar dosing regimen (drug infusion once every 7 days) ^22^. The reasons why sC5b-9 increases during eculizumab treatment, apparently diverging from clinical course, remain to be fully elucidated, but argue against using sC5b-9 as a reliable biomarker of disease activity in COVID-19.

Both inhibitors led to sustained inhibition of *ex vivo* AP-mediated complement hemolytic activity (AP50) in COVID-19 patient sera (Fig. 4, C). While AMY-101 treatment resulted in complete abrogation of AP activity throughout the treatment, a residual hemolytic activity (ranging between 7-11.5%) was detected in patient sera dosed with eculizumab on days 2 and 7 respectively (Fig 4, C). This ‘leakage’ in activity likely correlates with the rebound of terminal pathway activation products (sC5b-9) on day 7.

Our comparative study of the patients’ clinical response following complement modulation with two distinct inhibitory strategies revealed both common and divergent traits. Clearly both C3 and C5 inhibition led to a prominent and sustained anti-inflammatory response that likely mirrors the potential of both approaches to quench the proinflammatory actions of the C5a-C5aR1 axis ^10^. However, the tendency towards a steeper initial decline of LDH levels in the AMY-101 group may reflect a broader therapeutic effect of C3 inhibition on microvascular endothelial injury, aberrant pulmonary vascularization and lung damage, likely mediated by the blockage of the C3a-C3aR axis, the attenuation of C3 opsonization on injured endothelial or alveolar cells or the abrogation of tissue-injurious AP amplification ^3^. Of note, enhanced C3-mediated signaling has been implicated as an early driver of the host inflammatory response to SARS-CoV-2-infection. C3 expression showed robust transcriptional upregulation in both lung epithelial cells and nasopharyngeal swabs of COVID-19 patients ^9, 27^, while both C3aR and CD46 expression in the myeloid, lymphoid and lung epithelial compartments appear to correlate with disease severity in COVID-19 ^28^.

The prominent decrease of neutrophil counts in AMY-101-treated patients indicates that concomitant interception of C3a and C5a-triggered inflammation abrogates neutrophil recruitment, having important implications for long-term organ function. The concomitant use of corticosteroids (methyprednisolone, dexamethasone) in the Ecu-cohort may have skewed this response, affecting neutrophil turnover, migration between the circulation and tissues and vascular adhesion ^29^. The prolonged presence of high neutrophil numbers may entail long-term consequences in Ecu-patients that remain to be determined in follow-up studies. Given the ability of COVID-19 neutrophils to produce procoagulant TF-bearing NETs in the presence of intact C3 activation ^8^, future studies should address whether these high neutrophil counts under C5 blockade invoke long-term implications for organ function, despite recovery from SARS-CoV-2-induced pneumonia.

COVID-19 lymphopenia has been mainly linked to T-cell hyper-activation and/or depletion, likely mediated by increased IL-6 or TNF-signaling, enhanced recruitment of lymphocytes to the respiratory tract or increased adhesion to the vascular endothelium ^30^. Interception of C3 signaling with AMY-101 could reverse T cell depletion through the rapid lowering of the IL-6 inflammatory burden on peripheral lymphocytes. Furthermore, both C3a and C5a instruct the homing of activated T cells into inflamed tissues by altering endothelial adhesion molecules such as V-CAM-1 ^31^. By blocking both mediators, AMY-101 could likely exert a more profound inhibitory effect on the homing activity or pulmonary endothelial adhesion of COVID-19 lymphocytes. While other C3-independent mechanisms may influence lymphocyte recovery, this finding could have important implications in the context of developing a COVID-19-directed therapy.

The trend towards a greater transient increase of platelets in AMY-101 treated patients likely indicates a broader impact of C3 inhibition on platelet consumption and COVID19-thrombocytopenia. This effect may be related to mechanisms such as C3aR-dependent platelet adhesion to the vascular endothelium, C3a-driven thrombus formation via endothelial P-selectin upregulation, or C3-mediated opsonophagocytosis ^32, 33^.

Lung impairment and SARS-CoV-2 associated ARDS were similarly attenuated in both patient cohorts, reflecting a robust anti-inflammatory response by both inhibitors. AMY-101 treated patients showed consistent respiratory improvement without having received concomitant treatment with corticosteroids, as in the case of the Ecu-cohort. While the concomitant use of steroids in the most severe Ecu-patients may have led to synergistic effects in lung function improvement ^34^, the profound clinical gain observed under both inhibitory strategies paves the way to larger randomized trials that will formally benchmark the efficacy of these inhibitors in a well-controlled setting.

The persistently high C3a levels in the Ecu-treated patients confirmed that C5 blockade does not interfere with upstream C3 activation in COVID-19. A fully operative C3a-C3aR axis under C5 blockade could potentiate: i) monocyte/neutrophil recruitment to the infected lungs ii) cytokine release from macrophages and lymphocyte hyper-activation and iii) endothelial cell-platelet-neutrophil interactions promoting procoagulant responses, endothelialitis and TMA ^35,3^. This finding provides a mechanistic basis for a broader therapeutic effect of C3 inhibition in COVID-19 thromboinflammation. A trend towards lower median C3a levels in ecu-patients, on day 7 (approximately 35% decrease over baseline) (Fig 4, panel A), may reflect the cumulative therapeutic effect of eculizumab on vascular/organ injury which likely results in lower tissue damage-triggered complement activation in later stages.

CP and LP activity have been implicated in SARS-CoV-2 pathogenesis through the interaction of the heavily glycosylated N protein with MASP-2 ^11^ and the colocalization of viral S protein, immune complexes and C4d on COVID-19 erythrocytes ^36^. Moreover, the prominent presence of C4d and MASP-2 deposits in the microvascular endothelium of COVID-19 biopsies argues for a key role of these pathways ^4, 37^, To date, it remains debated whether the AP contributes to dysregulated complement activation in COVID-19. In our study, while plasma FB levels remained constant under C3 inhibition, they exhibited a consistent decline in all COVID-19 patients under eculizumab treatment, suggesting that in the presence of C5 blockade there is ongoing AP amplification (C3 convertase activity) that may lead to consumption of FB (Fig 4, D). To our knowledge, this is the first indication of ongoing AP activity in COVID-19 that is effectively blocked by AMY-101. Furthermore, these data are consistent with the ‘leakage’ in complement inhibition reflected by residual AP activity and rebound of sC5b-9 observed on day 7 from eculizumab treatment. The possibility that total FB levels are subject to the combined effect of protein consumption and altered biosynthesis due to an acute phase response associated with the viral infection cannot be excluded. Further studies interrogating the presence of activated FB fragments (Bb, Ba) in COVID-19 patients are expected to shed more light on the precise role of the AP. The notion that these reduced FB levels likely reflect AMY-101’s potency in blocking AP activity is further supported by a significant decrease of Bb and Ba fragments in AMY-101 treated patient samples (data not shown).

In conclusion, we have shown that clinical complement inhibition affords significant therapeutic benefit in COVID-19 patients by intercepting key SARS-CoV-2-induced thromboinflammatory pathways. This robust anti-inflammatory response culminates in respiratory improvement and resolution of COVID19-associated ARDS. C3 inhibition may exert a broader therapeutic effect in COVID19 patients by intercepting simultaneously upstream (C3-mediated) activation, AP amplification and terminal pathway activity, thereby preventing NET generation and subsequent thrombotic microangiopathies (supplemental Fig. 2). Signs of ongoing AP activation in COVID-19 patients under C5 blockade may point to an amplifying role of the AP in driving complement-mediated pathology, regardless of initiating route. Furthermore, the rebound of C5b-9 levels in these patients warrants further investigation as it may reflect unique pathogenic events leading to excessive C5 activation or drug dosing-related issues. Possible accumulation of sC5b-9 due to reduced clearance compared to smaller-sized fragments, or other yet poorly defined mechanisms, cannot be ruled out, indicating that C5b-9 may not be a reliable biomarker for monitoring ongoing complement activity in COVID-19 patients ^38^. The broader inhibitory profile of AMY-101 is associated with qualitative traits of improved clinical response over eculizumab. These improved clinical correlates indicate a broader engagement of C3-mediated pathways in COVID-19 pathophysiology. In light of the recent negative results from Phase III trials evaluating anti-IL-6 therapies in severe COVID-19 ^39^, complement inhibition emerges as a more comprehensive strategy to block IL-6 release and dampen the maladaptive host inflammatory response to SARS-CoV-2 ^5^. Future randomized controlled trials will conclusively discern the relative clinical efficacy of these two anti-complement strategies in COVID-19 patients.

## Data Availability

Data will be available upon request.

## Acknowledgments

The authors thank Quidel Corporation for the generous gift of its proprietary ELISA kits for the measurement of C3a AND sC5b-9 levels in all COVID-19 patients. This study was supported in part by the São Paulo Research Foundation (FAPESP) grant no. 13/08135-2, the Institutional National 5×1000 Grant (to F.C.) and the Dolce & Gabbana Fashion Firm (to C.G.). J.D.L. thanks Ralph and Sallie Weaver for the generous endowment of his professorship.

## Disclosure of Conflicts of Interest

JDL is the founder of Amyndas Pharmaceuticals which develops complement inhibitors for therapeutic purposes, and inventor of patents that describe the therapeutic use of complement inhibitors, some of which are developed by Amyndas. JDL is also the inventor of the compstatin technology licensed to Apellis Pharmaceuticals (i.e., 4(1MeW)7W/POT-4/APL-1 and PEGylated derivatives such as APL-2/pegcetacoplan and APL-9). A.M.R. has received research support from Alexion Pharmaceuticals, Novartis, Alnylam and Ra Pharma and lecture fees from Alexion, Novartis, Pfizer and Apellis, and served as member of advisory–investigator boards for Alexion, Roche, Achillion, Novartis, Apellis and Samsung, and as a consultant for Amyndas. B.N. is a shareholder and consultant in Tikomed and iCoat Medica. M.H.-L. holds a patent on compositions of matter and methods for the diagnosis and treatment of sepsis by C5a inhibitory strategies licensed to InflaRx. The other authors declare no competing interests.

## Supplemental data

### Supplemental methods

**a) Exclusion criteria** for the enrollment of COVID-19 patients in the Phase I/II trial of eculizumab were as follows:

Known hypersensitivity and / or previous eculizumab therapy; septic shock and / or multiple organ failure syndrome; history of infection by human immunodeficiency virus (HIV), hepatitis B virus (HBV) or hepatitis C (HCV) (with the exception of chronic infections treated or cured for HBV and HCV, which will be accepted); neoplasms in activity or under treatment, except for basal cell carcinomas; any major surgery, extensive radiation therapy, delayed toxicity chemotherapy, biological therapy, or immunotherapy within 6 weeks prior to clinical trial screening; any investigational medication other than the study drugs in the 6 weeks before the first dose of the study drug; history of chronic liver disease (Child-Pugh B or C); history of chronic kidney disease (glomerular filtration rate <30 mL / min / 1.73 m^2^); any contraindication to the use of penicillin for bacterial meningitis’s prophylaxis during the use of eculizumab; any contraindications to the use of contraceptive methods in childbearing-age women.

**b) Exclusion criteria** for enrolling COVID-19 patients in the AMY-101 compassionate use program at San Raffaele Hospital (Milan, Italy) were as follows: evidence of bacterial infection; concomitant administration of other immunosuppressive biologic agents.

Supplemental Tables

**Supplemental table S1.**
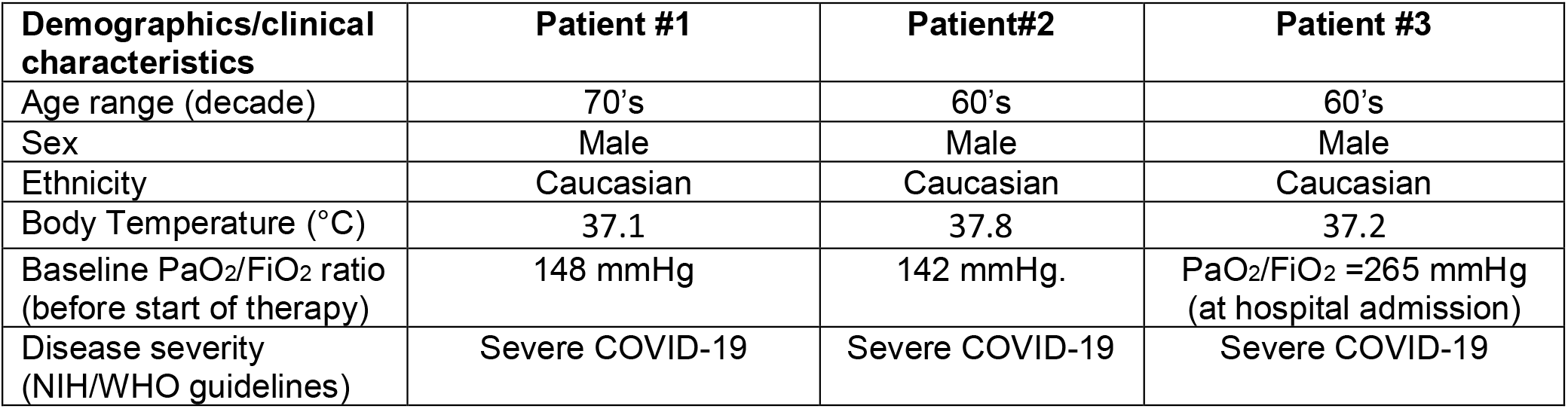
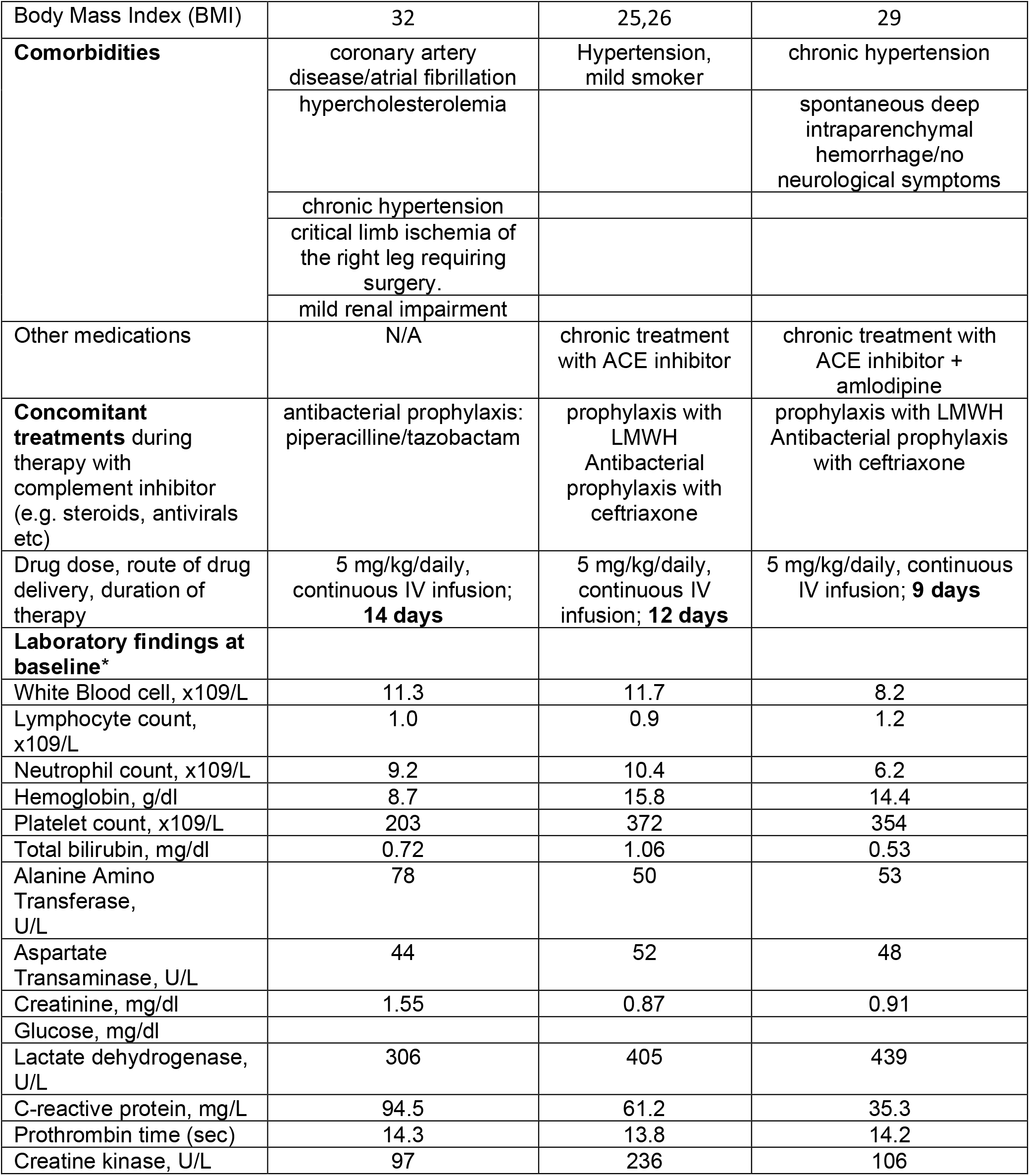
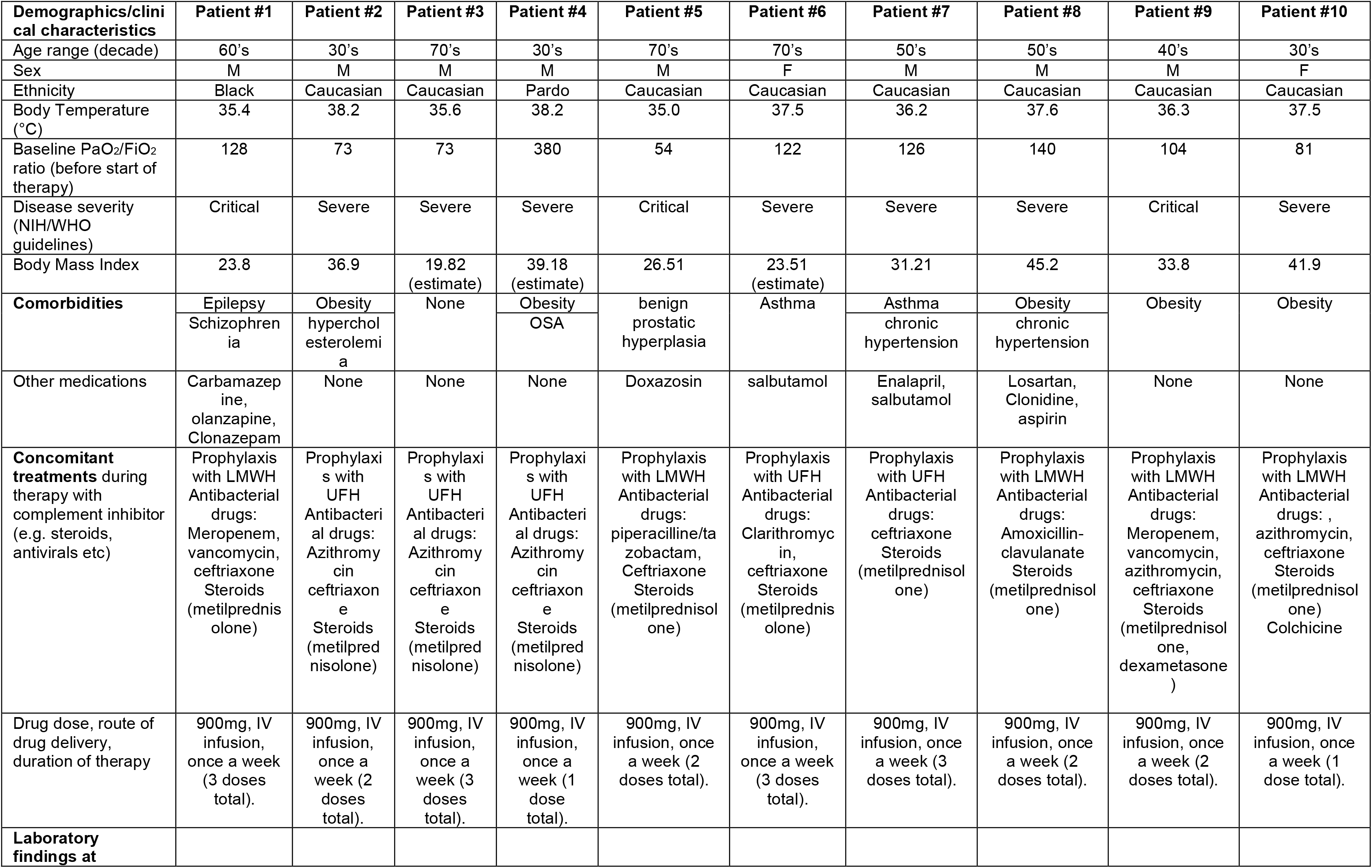
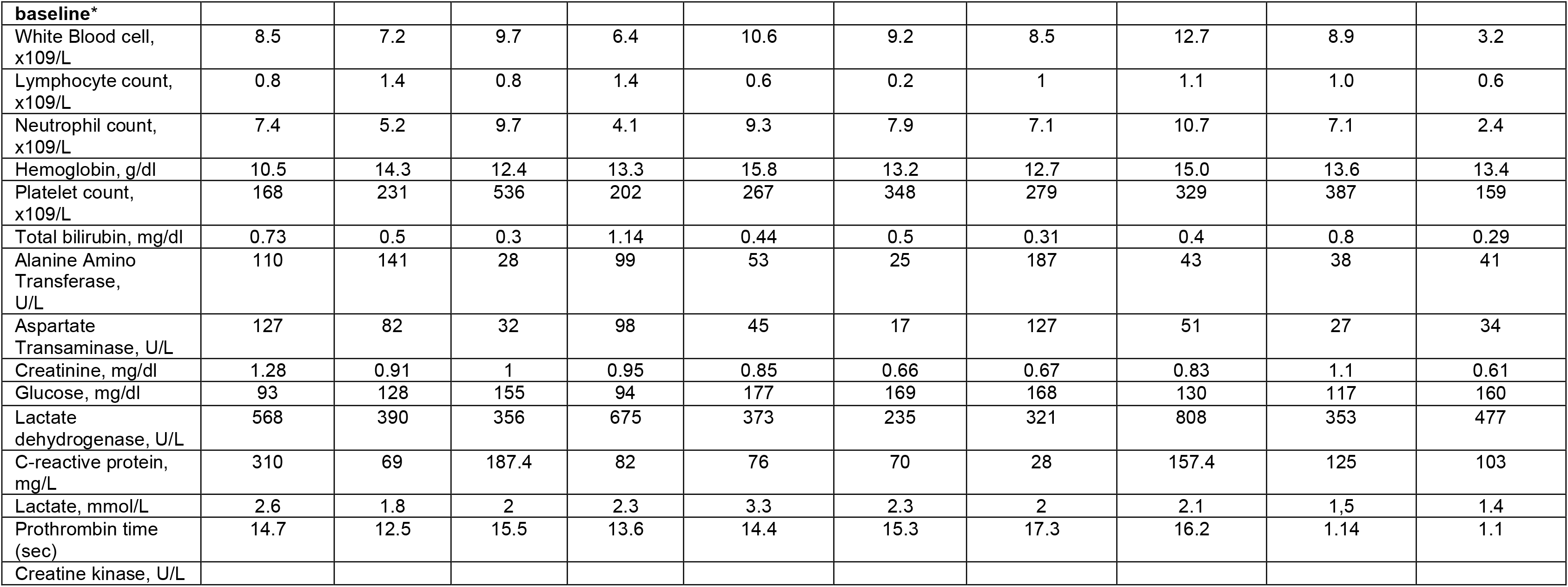
Patient demographics and clinical findings at baseline

**Supplemental Table S2.** Severe adverse events in the cohort of patients receiving eculizumab.

| Patient | Adverse Event | Grade (CTCAE) |
| --- | --- | --- |
| UPN1 | Disseminated intravascular coagulation | 3 |
| UPN1 | Anemia | 3 |
| UPN5 | Pulmonary thromboembolism | 3 |
| UPN6 | Myocarditis | 3 |

None of the severe adverse events were likely to be attributable to the study drug.

**Supplemental Figure 1:**
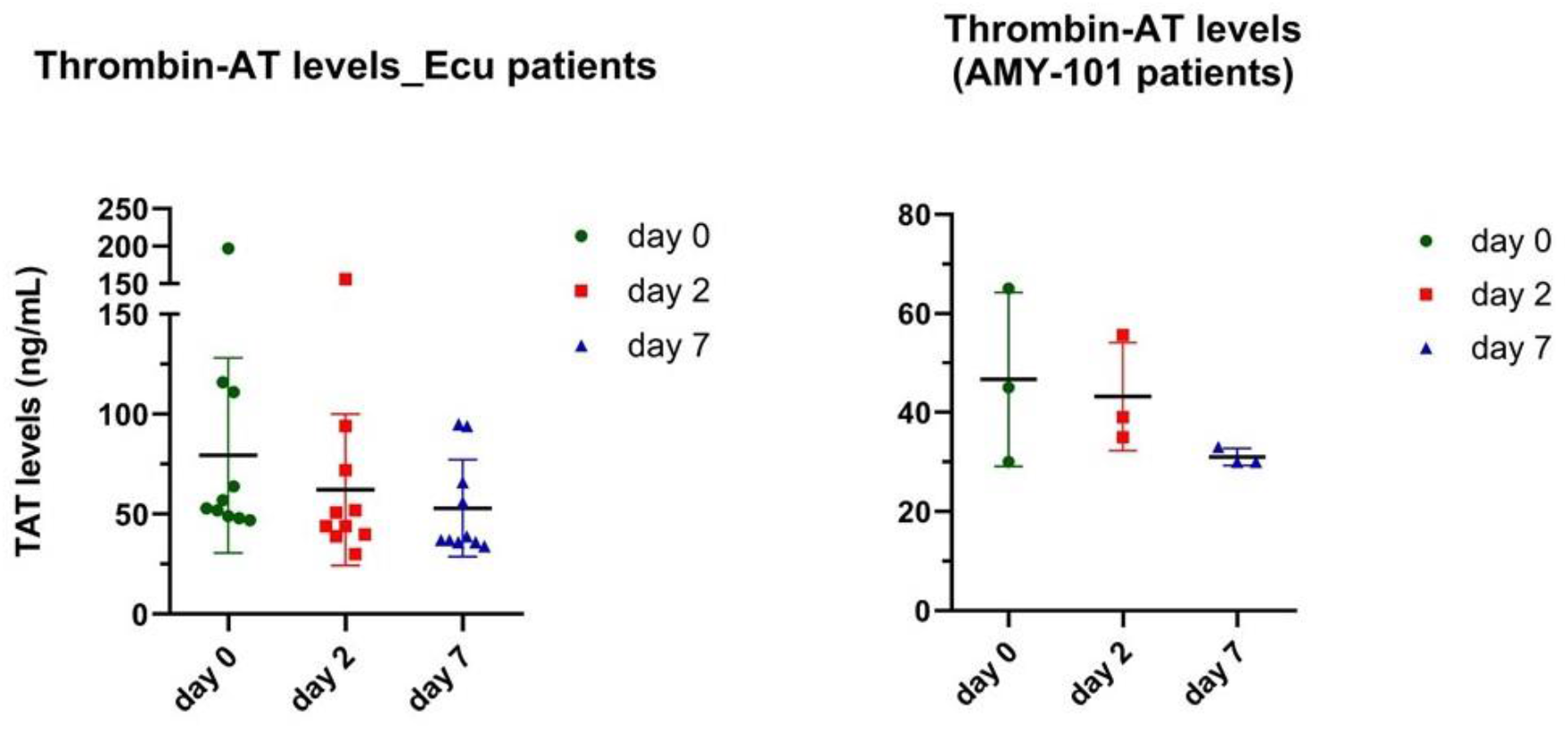
Both C3 and C5 inhibition impact thrombogenic pathways that promote thrombin generation in COVID-19. Complement inhibition resulted in reduction of thrombin-antithrombin complexes (TAT levels) in first 7 days of therapy. TAT complexes were measured in EDTA-plasma samples collected from COVID-19 patients dosed either with the C3 therapeutic AMY-101 (right panel) or with the C5-targeting mAb eculizumab (left panel). A similar reduction of TAT levels is observed in both patient groups from baseline through day 7, indicating a therapeutic effect of both inhibitors on COVID-19 associated coagulopathy. TAT complexes were quantified as described previously (

**Supplemental Figure 2.**
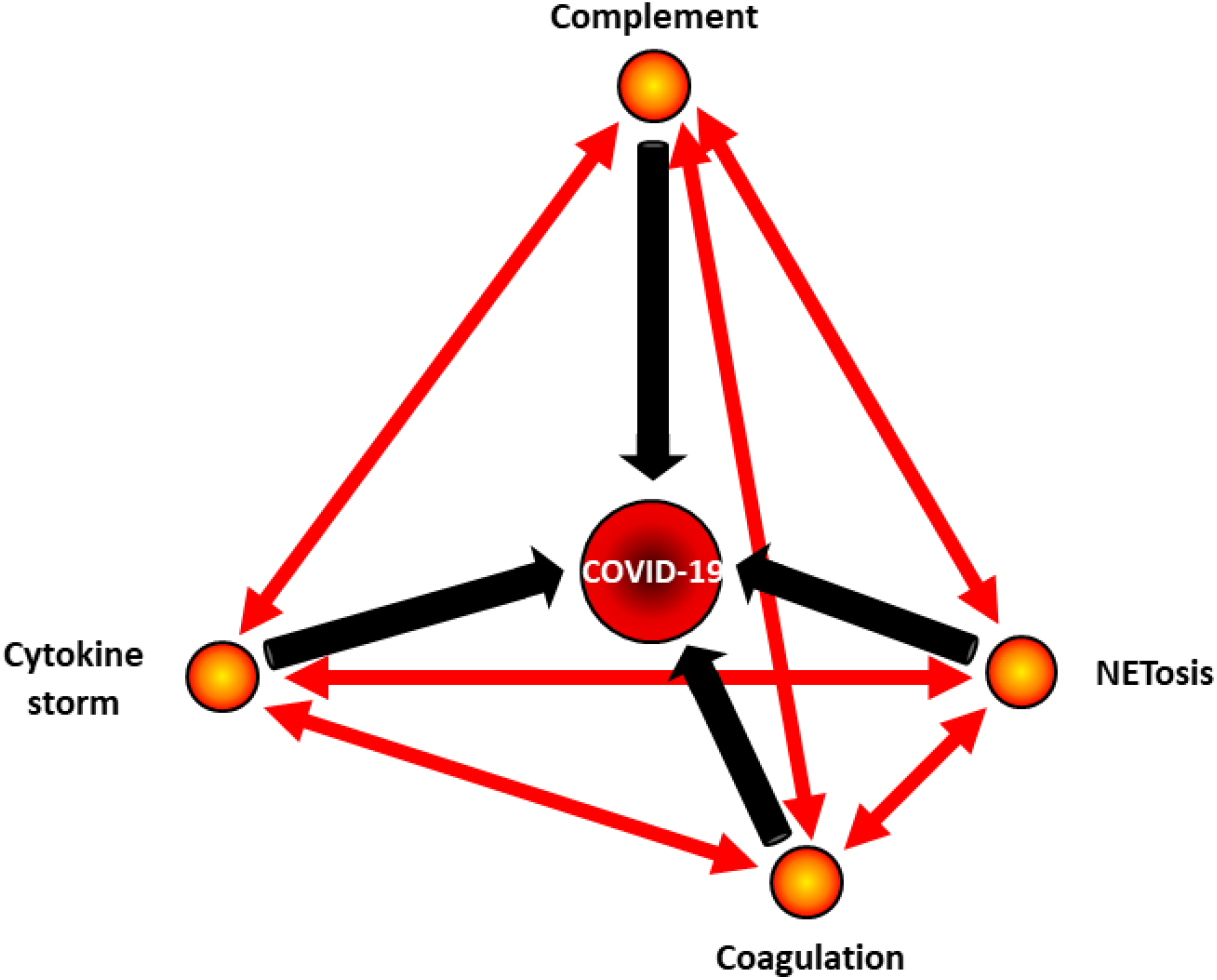
The tetrahedral pathophysiology of COVID-19. Complement activation plays a key role in the pathophysiology of COVID-19, eventually leading to the cytokine storm (associated with lung disease) and thrombophilia (accounting for multi-organ thrombotic microangiopathies). Thrombo-inflammation has emerged as a hallmark of COVID-19 immunopathology and neutrophil-driven NETosis appears to be a key disease-exacerbating mechanism cross-linked with all other pathogenic events. Indeed, complement activation may trigger NETs generation, which in turn amplifies complement activation, thereby enhancing inflammation and thrombophilia. Complement activation, cytokine-driven inflammation, NETosis and thrombophilia are closely embedded in the pathophysiology of COVID-19, and therapeutic interventions aiming to ‘defuse’ this detrimental loop need to interfere with early pathogenic events, such as proximal complement activation, preceding the tissue damage associated with thrombo-inflammation.

## Notes

### Clinical Trial

Study includes ten (10) consecutive COVID-19 patients enrolled in a phase I/II single arm clinical trial, coordinated by the University Hospital, University of Sao Paulo, Ribeirao Preto School of Medicine, Ribeirao Preto (Brazil) http://www.ensaiosclinicos.gov.br, RBR-876qb5 (RBR-876qb5

Eculizumab for the treatment of Covid-19 severe cases)

### Clinical Protocols

http://www.ensaiosclinicos.gov.br/rg/RBR-876qb5/

### Funding Statement

This study was supported in part by the Sao Paulo Research Foundation (FAPESP) grant no. 13/08135-2, the Institutional National 5×1000 Grant (to F.C.) and the Dolce & Gabbana Fashion Firm (to C.G.).

### Author Declarations

The phase I/II single arm clinical trial of eculizumab (http://www.ensaiosclinicos.gov.br, RBR-876qb5) was approved by Comissao Nacional de Etica em Pesquisa (CONEP), protocol number 30522020.0.0000.5440.

The compassionate use program of AMY-101 was approved by the IRB of San Raffaele Hospital, Milan, Italy (Approval March 26th 2020 by Ethical Committee of Istituto Nazionale per le Malattie Infettive Lazzaro Spallanzani I.R.C.C.S., Parere N. 35 del Registro delle Sperimentazioni).

